# Preterm birth, socioeconomic status, and white matter development across childhood

**DOI:** 10.1101/2025.06.10.25328994

**Authors:** Katie Mckinnon, Manuel Blesa Cábez, Melissa Thye, Selina Abel, Rebekah Smikle, Jean Skelton, Lorena Jiménez-Sánchez, Kadi Vaher, Gemma Sullivan, Amy Corrigan, Gayle Barclay, Charlotte Jardine, Iona Gerrish, Donna McIntyre, Yu Wei Chua, Ray Amir, Alan J. Quigley, Cheryl Battersby, Athanasios Tsanas, G David Batty, Rebecca M. Reynolds, Simon R. Cox, Heather C. Whalley, Michael J. Thrippleton, Mark E. Bastin, Hilary Richardson, James P. Boardman

## Abstract

**Background:** Preterm birth and socioeconomic status (SES) are associated with brain development in early life, but the contribution of each over time is uncertain. We examined the effects of gestational age (GA) and SES on white matter microstructure in the neonatal period and at five years.

**Methods:** Participants included preterm and term children. Diffusion MRI was collected at term-equivalent age (n=153 preterm, n=90 term [127/243 female]) and from a subset at five years (n=26 preterm, n=32 term [22/58 female]). We assessed linear associations of GA, SES (Scottish Index of Multiple Deprivation [SIMD] and maternal education), and GA×SES interactions on fractional anisotropy (FA) using tract-based spatial statistics. We compared the proportion of voxels with significant associations between timepoints.

**Results:** In preterm neonates, higher GA and higher maternal education, but not SIMD, were associated with higher FA (p corrected for family-wise error rate, p_FWER_<0.05). GA-FA associations depended on maternal education and SIMD (β=|0.001-0.005|, p<0.001). At five years, the strength and direction of GA-FA associations depended on SIMD (β=|0.013-0.028|, p<0.001), but not maternal education. In term infants, lower SES was associated with higher FA at the neonatal timepoint only (p_FWER_<0.05).

**Conclusions:** Preterm birth and SES both shape brain development at birth and continue to do so at five years. The SES measure most strongly associated with FA in preterm infants switches from a family-level (i.e. maternal education) to neighborhood-level (i.e. SIMD) measure between birth and five years, which suggests strategies to mitigate adverse effects of social inequalities on development may require adaptation as children grow.

## 1. Introduction

### 1.1 Preterm birth

Preterm birth affects 10% of children worldwide and is a leading cause of altered white matter development (encephalopathy of prematurity, EoP) and subsequent neurodevelopmental impairment (1,2). A consistent finding across diffusion magnetic resonance imaging (dMRI) studies is that, compared to term infants, preterm infants have microstructural differences in white matter, including decreased fractional anisotropy (FA) (3–5), which is a recognized marker of EoP (6,7). FA has proven useful for investigating upstream determinants of brain development; for example, it is associated with postnatal sepsis, histological chorioamnionitis (HCA), bronchopulmonary dysplasia (BPD), necrotizing enterocolitis (NEC), and suboptimal neonatal nutrition (3,8–13), and it associates with neurodevelopmental outcomes (14).

Meta-analytic evidence suggests that variations in brain morphology and microstructure after preterm birth persist (15). For example, studies using dMRI, magnetization transfer ratio, and T1-weighted/T2-weighted signal ratio suggest differences in preterm compared to term children at 4-7 years and in early adolescence (16–20). We have built a library of longitudinal brain image data linked to phenotypic data to investigate risk and resilience factors for healthy brain growth in preterm children (21,22).

### 1.2 Socioeconomic status

In the general population, brain structure, cognition, and educational attainment are associated with SES (23). A systematic review of neuroimaging studies across individuals aged 1-24 years reported associations between SES and structural and dMRI measures (24). The most consistent finding pertaining to diffusion measures in later childhood is positive higher SES-FA association across some, but not all, white matter tracts (24,25). However, in children under five years, data are inconsistent with studies reporting positive (26–30), negative (31,32), and no SES-FA associations (26,29–31,33,34).

We previously reported that SES and GA are associated with neonatal brain regional volumes, including white matter regions (35). Compared to term infants, there is a higher prevalence of socio-economic disadvantage in preterm infants (36,37), so these children are at risk of the effects of both preterm birth and low SES on brain development.

SES is a multifaceted construct and can be operationalized using neighborhood-level (e.g. postcode-derived deprivation indices) and family-level (e.g. parental education) measures (38,39). These measures, which are only weakly to moderately correlated with one another, capture different aspects of SES and have different impacts on regional brain volumes in preterm infants (35). We found more anatomically widespread associations between neonatal regional brain volumes with family-than neighborhood-level SES (35). In term infants, neighborhood measures may matter more than parental education as children age (73–76). The relationships between SES measures, GA, and white matter microstructure across childhood are unknown. Understanding these relationships is needed to target appropriate social support for vulnerable infants.

### 1.3 Aim and hypotheses

We aimed to investigate associations and interactions between SES, preterm birth, and white matter microstructure at term-equivalent age and five years by testing four hypotheses:

1. In preterm infants, lower SES and lower GA associate with lower FA across the white matter skeleton following mutual adjustment at the neonatal and five-year timepoints, and associations of preterm birth are stronger in children from lower SES;
2. SES-FA associations are partially attenuated when adjusting for preterm exposures (sepsis, HCA, BPD, NEC, and low breast milk intake) at the neonatal and five-year timepoints;
3. In term infants, lower SES is associated with lower FA across the white matter skeleton at the neonatal and five-year timepoints;
4. In all participants, associations between neighborhood-level measures of SES and FA in white matter are more widespread at five years than in the neonatal period.

## 2. Methods and materials

### 2.1 Participants

Participants were preterm (<33 weeks’ GA) and term infants born at the Royal Infirmary Edinburgh, UK, and recruited into the Theirworld Edinburgh Birth Cohort (TEBC). This longitudinal study was designed to investigate the effect of preterm birth on brain development and long-term outcomes (21,22). Recruitment was between 2016 and 2021. Exclusion criteria were major congenital malformation, chromosomal abnormality, and major parenchymal lesions. Ethical approval was from the National Research Ethics Service, South-East Scotland Research Ethics Committee (REC 16/SS/0154), and NHS Lothian Research and Development (2016/0255). Parents provided written informed consent at both timepoints.

At term-equivalent age, 153 preterm and 90 term children had dMRI included (Table 1). Five-year dMRI was included for 26 preterm and 32 term children (Table 1). Of these, 52 children were included at both timepoints (24 preterm and 28 term children). There was no difference in sex distribution between groups (Table 1). Parent-reported child ethnicity did not differ between groups (Table 1) and was representative of Edinburgh (40). Figure S1 gives a flowchart of participants.

**Table 1.**
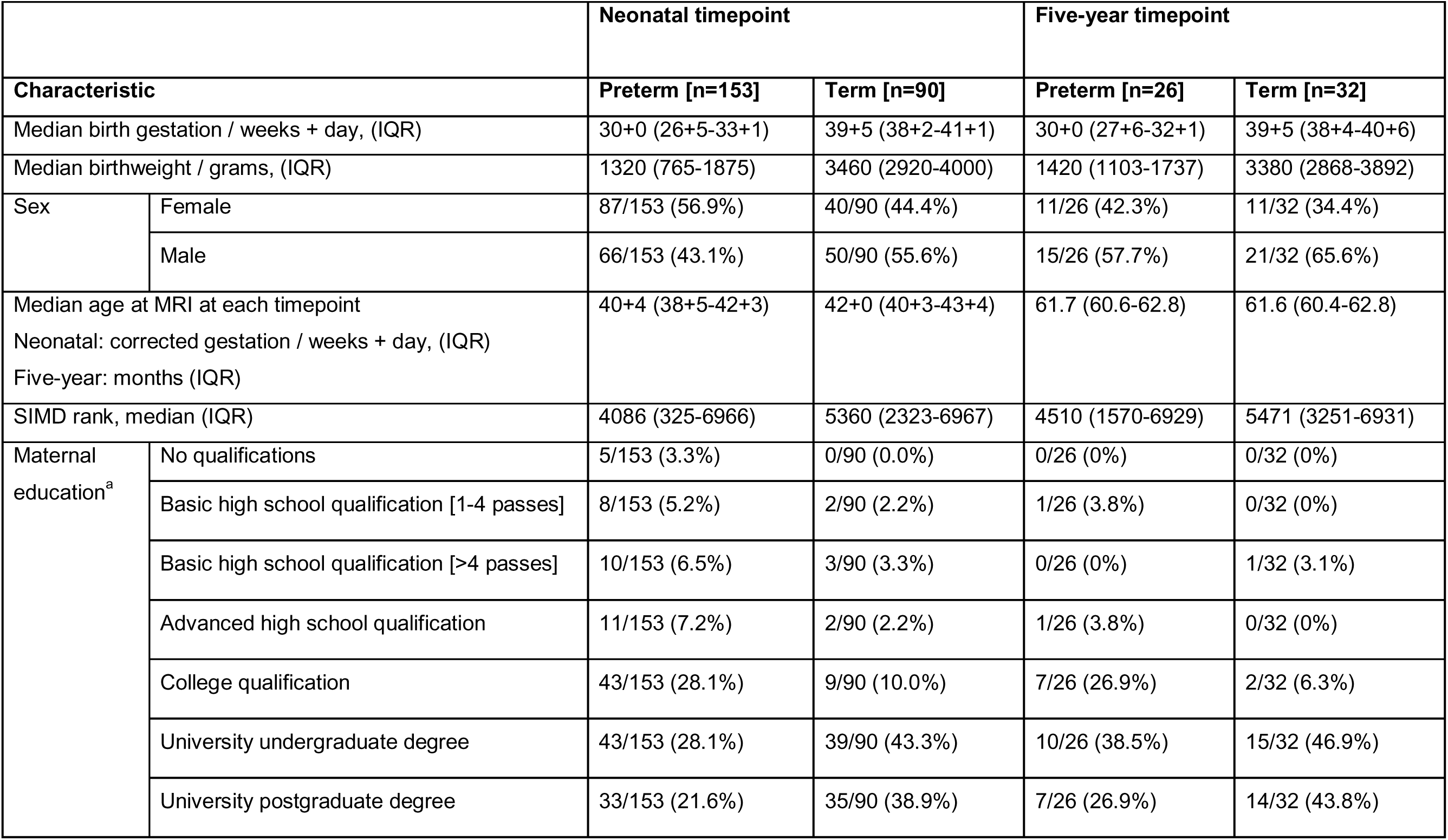
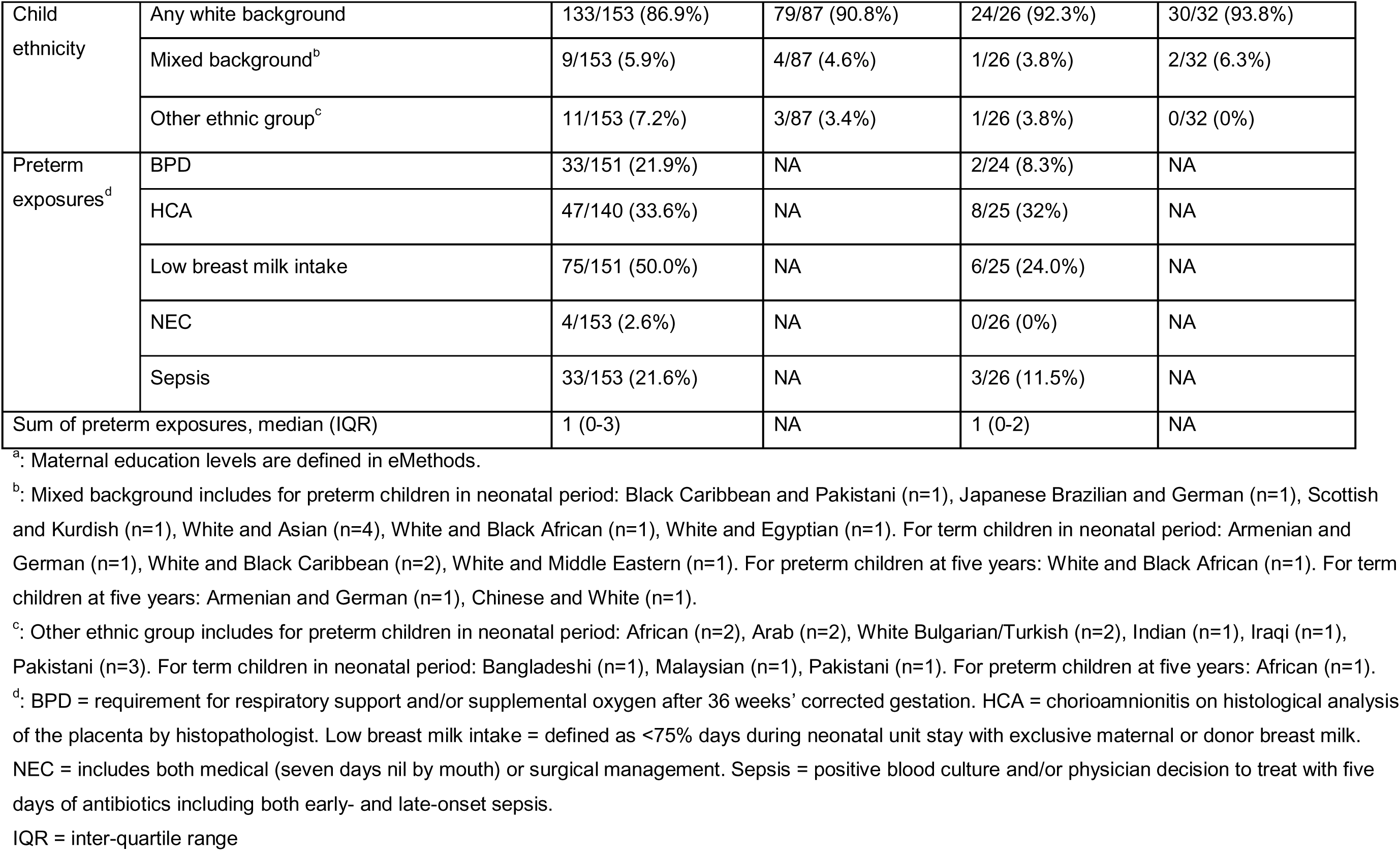
Participant characteristics.

### 2.2 Imaging

#### 2.2.1 Magnetic resonance imaging acquisition

At term-equivalent age, MRI scans were performed according to published protocol (21). A Siemens MAGNETOM Prisma 3T MRI clinical scanner (Siemens Healthcare, Erlangen, Germany) and 16-channel phased-array pediatric head receive coil were used to acquire a multishell axial dMRI scan (16×b=0s/mm^2^, 3×b=200s/mm^2^, 6×b=500s/mm^2^, 64×b=750s/mm^2^, 64×b=2500s/mm^2^) with optimal angular coverage (see eMethods for full protocol). Infants were fed, wrapped, and slept naturally. Flexible earplugs and neonatal earmuffs (MiniMuffs, Natus) were used for acoustic protection. Infants were monitored throughout, and scans were supervised by a doctor or nurse trained in neonatal resuscitation.

At five years, the same MRI scanner with a 32-channel phased-array head receive coil was used to acquire a 3D T1-weighted MPRAGE scan (voxel size=1mm isotropic) and a multishell axial dMRI scan (15×b=0s/mm^2^, 3×b=200s/mm^2^, 6×b=500s/mm^2^, 64×b=1000s/mm^2^, 64×b=2000s/mm^2^) with optimal angular coverage (see eMethods for full protocol) (22). Children were introduced to the scanner using a child-friendly booklet and mock-scan session with simulated MRI scanner. Children wore in-ear headphones (Nordic Neuro Lab, Bergen, Norway) to minimize scanner noise and enable listening to movies of their choice. During the scan, a researcher stood at the foot of the bore to monitor the child. If the child moved visibly, the researcher reminded them to stay still.

All structural images were reviewed by a radiologist with experience in pediatric MRI (AJQ) and conventional findings from the neonatal timepoint are reported (41). No participants in this study had major parenchymal lesions (Figure S1).

#### 2.2.2 Data pre-processing

At term-equivalent age, raw images were visually inspected before processing and low-quality images discarded (Figure S1). For each participant, the two dMRI acquisitions were concatenated and denoised using Marchenko-Pastur principal component analysis-based algorithm with MRtrix3 (42,43); eddy current, head movement and echo planar imaging (EPI) geometric distortions were corrected using outlier replacement and slice-to-volume registration (44–47) using FMRIB Software Library (FSL) (48); bias field inhomogeneity correction was performed by calculating the bias field of mean b0 volume and applying correction to volumes (49) using MRtrix3.

At five years, pre-processing was similar. Images were visually inspected, b=200s/mm^2^ removed, dMRI data denoised (42,43) and Gibbs unringing performed using MRtrix3 (50). Eddy current, head movement and EPI geometric distortions were corrected using outlier replacement and slice-to-volume registration incorporating susceptibility-by-movement interactions with eddy currents (44–47,51); bias field inhomogeneity correction was performed by calculating the bias field of mean b0 volume and applying correction to volumes (49). Finally, images were resampled to 1.5mm isotropic voxels.

Diffusion tensor imaging (DTI) maps were calculated using dMRI processed images to obtain FA. A DTI model was fitted in each voxel using weighted least-squares method DTIFIT in FSL, using only b=750s/mm^2^ shell for neonates and b=1000s/mm^2^ for five years.

#### 2.2.3 Voxel-wise analyses

Voxel-wise statistical analyses of FA data were carried out using Tract-Based Spatial Statistics (TBSS) (52) and Randomise in FSL (48). Separately for the two timepoints, all participants’ FA data were transformed into a common space, i.e. the most representative participant for each timepoint, identified as the participant requiring minimum transformation. This used affine and non-linear registration (53,54), with additional initial rigid registration for term-equivalent age (10). In addition, the selected participant was non-rigidly registered to age-specific templates for each timepoint (55–58).

This transformation was concatenated to the previous and applied to each participant. Next, the mean FA image was created separately for timepoints and thinned to create a mean FA skeleton representing the centers of all tracts common to the group, with threshold FA 0.15 for term-equivalent age, and 0.2 for five years. Each participant’s aligned FA data was projected onto their corresponding skeleton and the resulting data fed into voxel-wise cross-participant statistics.

### 2.3 Variables and covariates

We assessed SES using two measures. Neighborhood-level SES was defined using the Scottish Index of Multiple Deprivation rank (SIMD, 2016) (59), derived from family postcode at birth, analyzed as a continuous measure from 1 (most deprived) to 6976 (least deprived). The index incorporates neighborhood indicators of income, employment, education, health, access to services, crime, and housing (59). For visualization of interaction effects, these were combined into two groups, namely SIMD quintiles 1-4 and SIMD quintile 5. Family-level SES was operationalized using ordered categories of maternal education, obtained from parent-reported highest educational qualification at recruitment (see supplementary eMethods for categories). For visualization of interaction effects, these were combined into two groups, namely maternal non-university education and university education.

GA was derived from medical records, as a continuous variable (60,61). Age at MRI scan was included as a continuous measure in all analyses for each timepoint. At term-equivalent age this was corrected GA in weeks, and at five years this was chronological age in years.

Sex was derived from medical records and included because there are sex differences in white matter development through childhood (62). Preterm exposures associated with FA were combined into a single measure with a total score of 0-5 (3,63), counting the following exposures for every infant: BPD, HCA, low breast milk intake, NEC, and sepsis (3,8–13). Preterm exposure definitions are detailed in eMethods.

### 2.4 Statistical analyses

#### 2.4.1 Tract-based spatial statistics

Voxel-wise statistical analysis of FA data was performed using Randomise in FSL (48,64). In timepoint-specific analyses, multivariable linear regression models assessed the relationship between FA and GA, SES (maternal education and SIMD separately), and an multiplication interaction between SES and GA (removed if no significant association seen), with sex, age at scan, and an intercept as covariates. For preterm infants, analyses were then adjusted for preterm exposures by inclusion as an additional covariate. Each analysis included family-wise error correction for multiple comparisons and threshold-free cluster enhancement, with statistical significance of *p* corrected for family-wise error rate (*p_FWER_*)<0.05 (65,66).

Statistical analyses were preregistered (67). In the whole cohort, in the neonatal period we found positive and negative associations between GA and FA, and small FA regions associated with an interaction between maternal education and GA (Tables S1-2, Figures S2-3). The interaction was such that higher GA associated with lower FA for children of mothers with no educational qualifications, but higher GA associated with higher FA for children of mothers with any educational qualifications (Figure S3). At the five-year timepoint, we found small regions with a positive association between GA and FA. GA and SIMD interacted, such that higher GA associated with lower FA for children in families living in the least deprived neighborhoods (Figure S2 and S4). These observations suggested that relationships between GA, SES and FA differ by gestational category.

Therefore, we undertook the voxel-wise statistical analyses separately for the preterm and term infants. Details of each regression model are in Table S3.

#### 2.4.2 Comparison of SES effects on FA between birth and five years

We counted the number of voxels where there was a statistically significant association (*p_FWER_*<0.05) between FA and GA, maternal education, SIMD, or a multiplication interaction between GA and each SES measure. We counted the total voxels tested at each timepoint to account for volume differences. We compared the proportion of the white matter skeleton with significant associations with each SES measure and each interaction between timepoints using McNemar tests in R.

## 3. Results

### 3.1 Associations between gestational age, socioeconomic status, and fractional anisotropy in preterm infants

#### 3.1.1 Neonatal timepoint

Among preterm infants at term-equivalent age (n=153), infants born at higher GA had higher FA in the corpus callosum, posterior corona radiata, and cingulum bundles, after adjustment for sex and age at scan, irrespective of SES measure included in the model. Figure 1A shows results adjusted for maternal education and similar results were seen in the model adjusted for SIMD.

**Figure 1.**
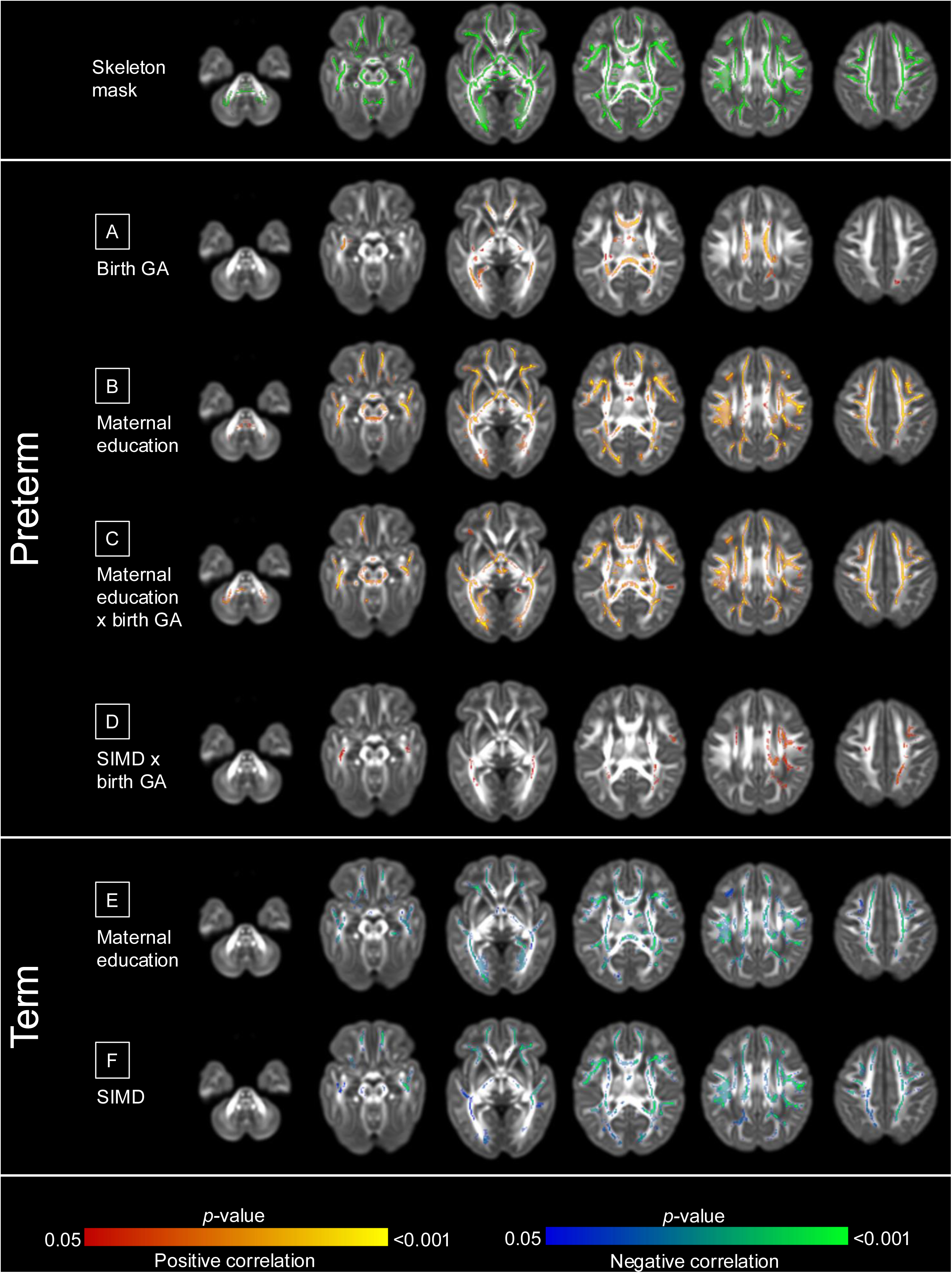
Voxels where fractional anisotropy associations were found in the neonatal period. In preterm infants (n=153), models were mutually adjusted for GA, SES (maternal education and SIMD separately), GA × SES interaction, sex, and age at MRI. A: GA, B: maternal education, C: maternal education × GA, D: SIMD × GA. In term infants (n=90), models were adjusted for sex and age at MRI. E: maternal education, F: SIMD. Results are reported after 5000 permutations, *p*-values corrected using threshold-free cluster enhancement and family-wise error correction with a significance level of *p*<0.05. For visualization: anatomic left is on the left side of the image. Red-yellow indicates a positive association, blue indicates a negative association. Results are displayed overlaid on the dHCP neonatal template (57) for the neonatal timepoint.

Higher maternal education was associated with higher FA across the white matter (Figure 1B), and there were anatomically widespread GA × maternal education interaction effects on FA (Figure 1C). Specifically, higher GA associated with higher FA for neonates of mothers with university education (n=76), whereas higher GA associated with lower FA for infants of mothers without university education (n=77) (β=0.003 and β=-0.002 respectively, *p*<0.001) (Figure 2A).

**Figure 2.**
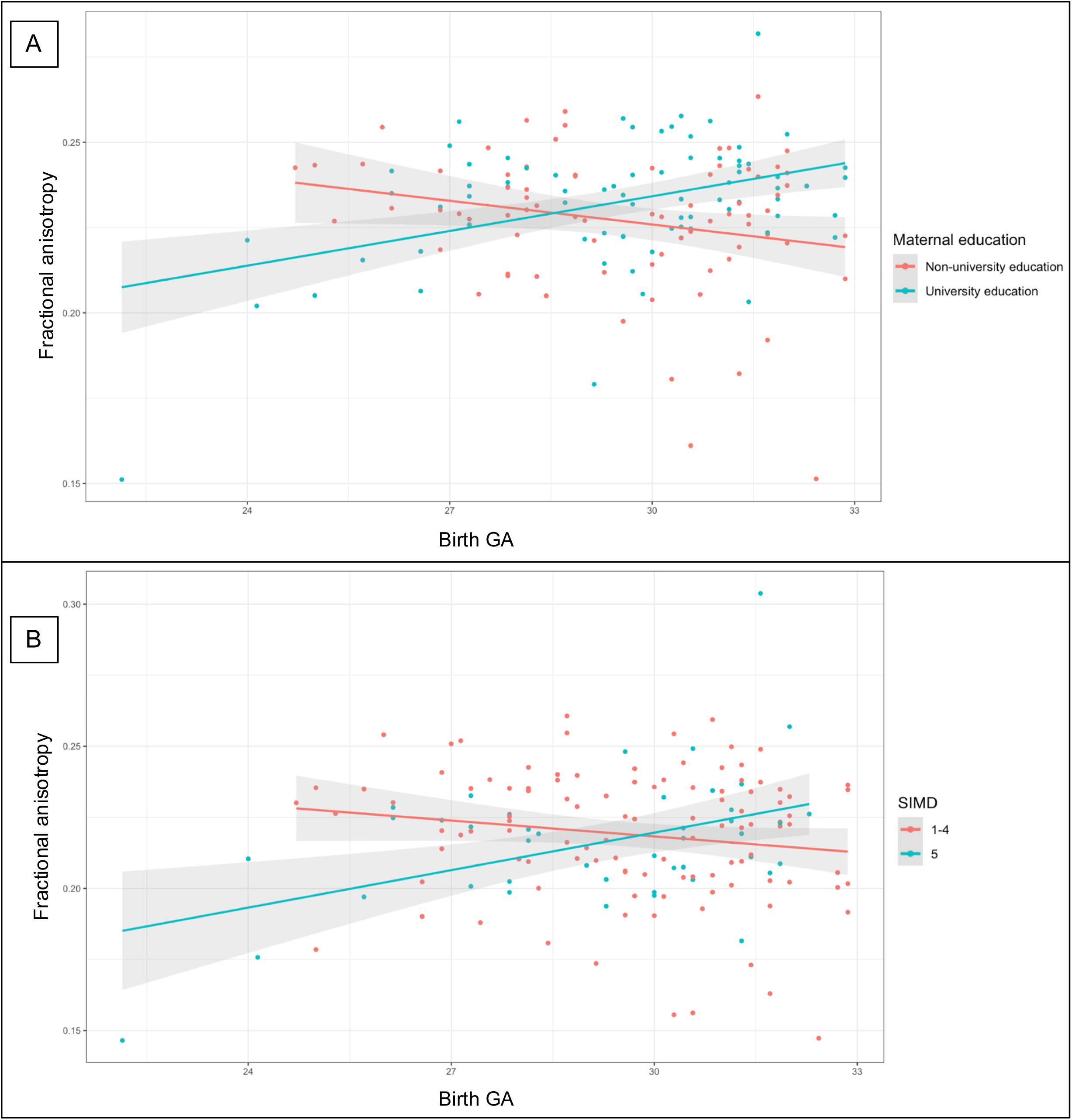
Interaction effects between gestational age and socioeconomic status on fractional anisotropy at term-equivalent age in preterm infants. A: Interaction effect between GA and maternal education on FA. B: Interaction effect between GA and SIMD on FA. Term-equivalent age in preterm infants (n=153). For each individual, mean FA was extracted from voxels where there are significant interaction effects. Interaction effects all *p*<0.001. Maternal education analyses were performed using maternal education as seven categories (see eMethods), visualized as linear regression lines for two groups for ease of interpretation, namely maternal non-university education (no qualifications, basic or advanced high school qualifications, or college qualification, n=77) and university education (undergraduate or postgraduate degrees, n=76). SIMD analyses were performed using SIMD rank as a continuous measure, visualized as two groups for ease of interpretation, namely SIMD quintiles 1-4 (n=109) and SIMD quintile 5 (n=44).

There were no independent associations between SIMD and FA at term-equivalent age. However, there was significant interaction between GA and SIMD with FA such that higher GA associated with higher FA in the corona radiata and superior longitudinal fasciculus, for children born into the least deprived neighborhoods (Figure 1D). The interaction is illustrated in Figure 2B, which shows higher GA is associated with higher FA for children whose families are living in less deprived neighborhoods (SIMD quintile 5, n=44), whereas higher GA associated with lower FA for children whose families are living in the more deprived neighborhoods (SIMD quintiles 1-4, n=109) (β=0.005 and β=-0.001 respectively, *p*<0.001).

Associations between GA and FA were no longer significant in models adjusted for co-morbidities of preterm birth (Table S4). However, associations between FA and maternal education, and the interaction between GA and maternal education or SIMD, were minimally affected (Figure 3A-C, Table S4).

**Figure 3.**
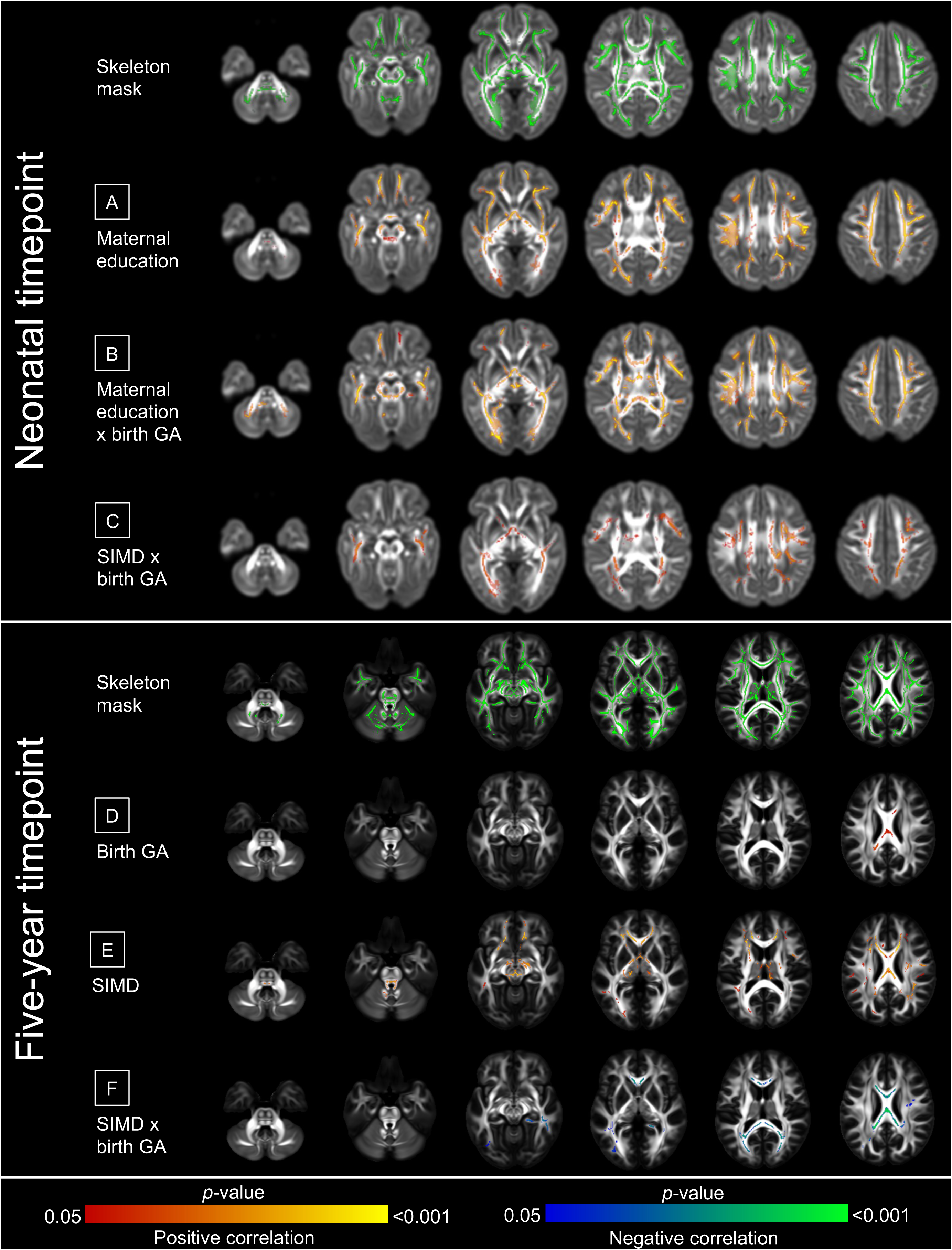
Associations between gestational age, socioeconomic status, and fractional anisotropy in preterm infants after adjustment for co-exposures of preterm birth. Voxels where FA associations were found in preterm infants at the neonatal timepoint (n=153) and at the five-year timepoint (n=26) after adjustment for co-exposures. Models were mutually adjusted for GA, SES (maternal education and SIMD separately), GA × SES interaction, sex, age at MRI, and sum of preterm exposures (BPD, HCA, low breast milk intake, NEC, and sepsis). In the neonatal period - A: Maternal education, B: maternal education × GA, C: SIMD × GA. At five years - D: GA, E: SIMD, F: SIMD × GA. Results are reported after 5000 permutations, p-values corrected using threshold-free cluster enhancement and family-wise error correction with a significance level of *p*<0.05. For visualization: anatomic left is on the left side of the image. Red-yellow indicates a positive association, blue indicates a negative association. Overlaid on the developing Human Connectome Project (dHCP) neonatal template (57) for the neonatal timepoint, and on the FSL HCP1065 (Human Connectome Project) adult template (56) for the five-year timepoint.

#### 3.1.2 Five-year timepoint

In preterm infants at five years (n=26), there were associations between higher GA and higher FA in the corpus callosum (Figure 4A), and between higher SIMD and higher FA in the corpus callosum, internal capsule, and brainstem (Figure 4B). There were associations in the corpus callosum between the interaction of GA × SIMD on FA, such that higher GA associated with higher FA in children living in more deprived neighborhoods, whereas higher GA associated with lower FA for children in less deprived neighborhoods (β=0.028 and β=-0.013 respectively, *p*<0.001, Figure 4C and Figure S5).

**Figure 4.**
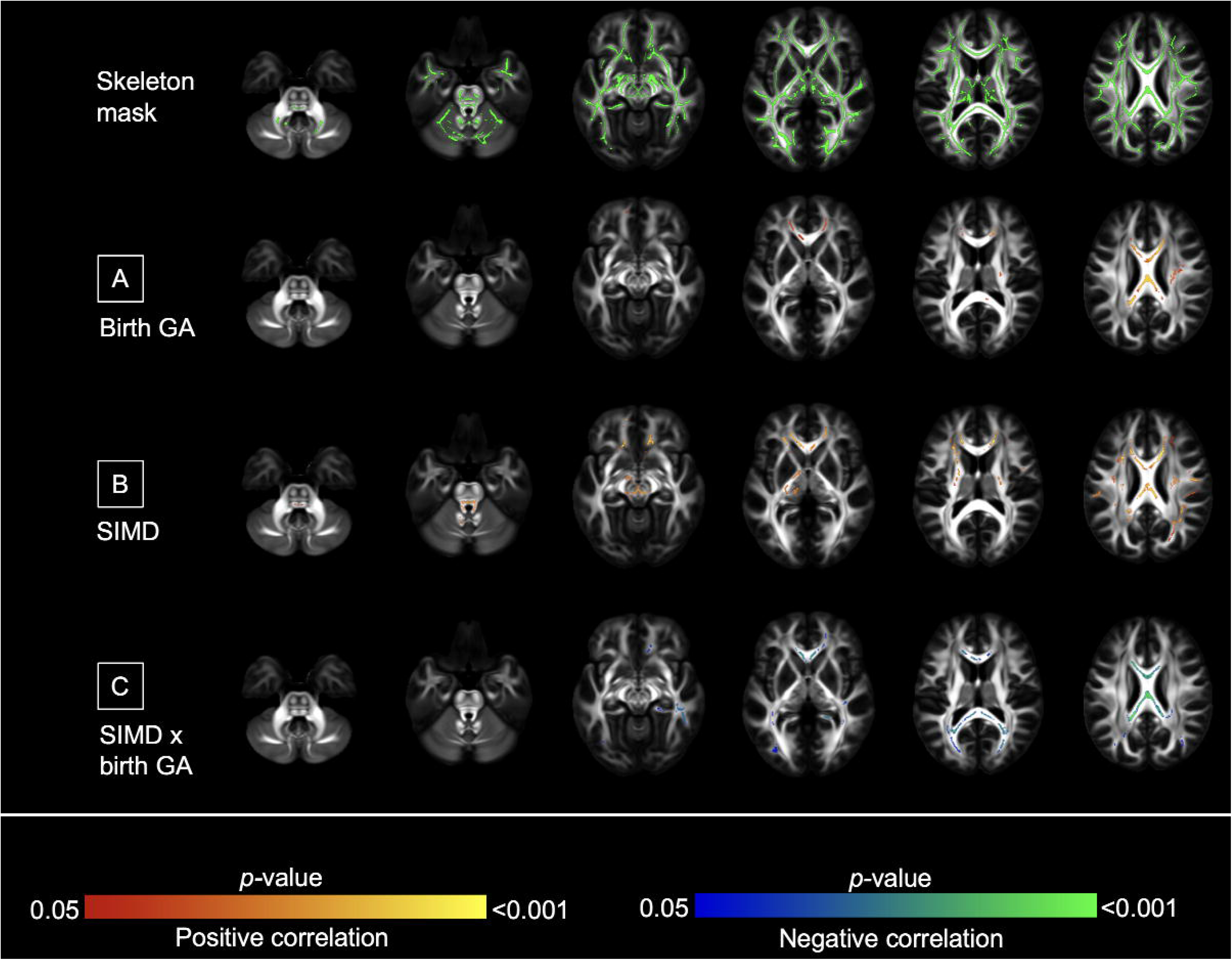
Voxels where fractional anisotropy associations were found at five years of age in preterm infants. Voxels where FA associations were found in preterm infants at the five-year timepoint (n=26). Models were mutually adjusted for GA, SES (maternal education and SIMD separately), GA × SES interaction, sex, age at MRI. A: GA, B: SIMD, C: SIMD × GA. Results are reported after 5000 permutations, *p*-values corrected using threshold-free cluster enhancement and family-wise error correction with a significance level of *p*<0.05. For visualization: anatomic left is on the left side of the image. Red-yellow indicates a positive association, blue indicates a negative association. Overlaid on the FSL HCP1065 adult template (56) for the five-year timepoint.

These remained significant after adjustment for perinatal exposures, although the proportion of the white matter skeleton associated with GA reduced (5.6% [5816/104030 voxels] to 0.9% [959/104030 voxels]) (Figure 3D-F, Table S4). There were no associations between FA and maternal education, or an interaction between GA and maternal education, at five years.

### 3.2 Associations between socioeconomic status and fractional anisotropy in term infants

#### 3.2.1 Neonatal timepoint

In term infants (n=90), higher SES associated with lower FA across the white matter (Figure 1E-F). Both maternal education (Figure 1E) and SIMD (Figure 1F) showed similarly anatomically widespread associations in term neonates, in contrast to preterm infants when maternal education has more widespread associations than neighborhood deprivation.

#### 3.2.2 Five-year timepoint

In term infants at five years (n=32), there were no associations between either SES measure and FA.

### 3.3 Differences in socioeconomic status-fractional anisotropy associations between the neonatal period and five years

In preterm infants, there were differences over time in the proportion of the white matter skeleton containing significant voxel-wise associations between SES and SES × GA interactions with FA (*p*<0.001). At the neonatal timepoint, maternal education (57.5%, 16890/29354 voxels) and GA × maternal education (53%, 15544/29354) were associated with FA in a higher proportion of the white matter skeleton compared to SIMD (0%) or an interaction between SIMD and GA (8.6%, 2526/29354). However, at five years, SIMD (14.8%, 15381/104030) or the interaction between GA and SIMD (6.7%, 7008/104030) had the most widespread associations with FA (Table 2).

**Table 2.** Number of significant voxels where FA associates with SES, GA, or an interaction between SES and GA.

| Group | Variable | Number (proportion) of significant voxels at the neonatal timepoint | Number (proportion) of significant voxels at the five-year timepoint |
| --- | --- | --- | --- |
| Preterm | GA (in model with maternal education) | 3934 (13.4%) | 0 |
|  | GA (in model with SIMD) | 4014 (13.7%) | 5816 (5.6%) |
|  | Maternal education | 16890 (57.5%) | 0 |
|  | Maternal education × GA | 15544 (53%) | 0 |
|  | SIMD | 0 | 15381 (14.8%) |
|  | SIMD × GA | 2526 (8.6%) | 7008 (6.7%) |
| Term | Maternal education | 12941 (44.1%) | 0 |
|  | SIMD | 12945 (44.1%) | 0 |

Number of significant voxels ( $p$ corrected for family-wise error rate ( $p_{FWE}$ )<0.05) where FA associates with SES, GA, or an interaction between SES and GA. Total number of voxels at neonatal timepoint = 29354, and at five years of age = 104030.

In term infants at both timepoints there were similar proportions of white matter showing SES-FA associations with each SES measure. At the neonatal timepoint, maternal education and SIMD were similarly associated with FA (44.1% [12941/29354] and 44.1% [12945/29354] voxels respectively). At five years, there were no associations between maternal education or SIMD with FA (Table 2).

## 4. Discussion

### 4.1 Key findings

In a contemporary birth cohort enriched for prematurity, GA and SES were associated with FA across the white matter skeleton during the neonatal period and at five years. The extent and direction of these associations depend on gestational category (preterm versus term) and SES measure.

Among preterm infants, the observation that higher GA is associated with higher FA within the white matter is consistent across many studies (7). Here we further show that GA-FA associations reduced after adjustment for preterm adverse co-exposures (sepsis, HCA, BPD, NEC, and low breast milk intake). This corroborates growing evidence indicating that it is not low GA *per se* that drives the development of encephalopathy of prematurity and adverse outcomes, but rather, it is adverse medical and environmental risks experienced by preterm infants, such as immune dysregulation and sub-optimal nutrition, that play a key role in atypical brain development of preterm infants (14,68,69). In addition, we found striking associations between SES and FA that were more widespread across the neonatal WM skeleton than those attributable to low GA. Of SES measures tested, maternal education was more strongly associated with neonatal brain development than neighborhood deprivation revealing a consistent pattern that for children from higher SES families, higher GA is associated with higher FA but in children from lower SES families higher GA associates with lower FA. In sum, the relationship between degree of prematurity and neonatal white matter development is modified by maternal education level and neighborhood deprivation.

There were differences in the direction of SES-FA associations between preterm and term infants. In preterm infants, higher maternal education was associated with higher FA, whereas in term infants, higher maternal education and living in a less deprived neighborhood associated with lower FA. This unexpected finding could be due to the greater susceptibility of the preterm brain to socioeconomic influences. Preterm infants may have an accelerated maturation of white matter, manifesting as higher FA, which is not seen in term infants. SIMD and maternal education appear to have similar effects in term infants, but among preterm infants, maternal education has a stronger role in the neonatal period. This could be due to the family-level measure better capturing adversities in the antenatal and early neonatal periods.

In preterm infants, when adjusting for preterm co-exposures and co-morbidities, associations between SES and SES × GA and FA are minimally changed, both in the magnitude of association and distribution of nominally significant voxels. This suggests that associations between SES and white matter microstructure are independent of these early life exposures. This may be because preterm co-exposures have such a strong influence on FA that this is independent of SES. However, interventions beyond the medical care of preterm infants are likely needed to effectively address the influence of SES on brain development.

In five-year-old preterm children, FA associated with SIMD and an interaction between GA and SIMD, indicating that for those with lower SES, higher GA was associated with higher FA, but there was a negative association between GA and FA in the higher SES group. In contrast, in term children at five years, SES measures were not associated with FA.

The finding of different directions of SES-FA associations at the two timepoints is consistent with the protracted development hypothesis (25); this could explain the negative SES-FA association in term infants and the negative GA-FA association for preterm infants from lower SES backgrounds in the neonatal period. This hypothesis describes how higher SES is associated with more protracted structural and functional development, with slower initial development followed by rapid development in later childhood and adolescence (25). Although reviews in older children have suggested a positive SES-FA association (24,25), the variability in findings in younger children (26,27,29–34) may be explained by a more complex relationship with age. These complex relationships are also consistent with previous morphometric similarity network analyses that demonstrate dysmaturation such that some networks have accelerated maturation while others are delayed (70). Given the varying nature and direction of SES-FA associations seen here, imaging techniques allowing more inference about microstructural development than FA, and those combining different microstructural indices are likely to be informative in future studies of SES-brain interactions in early life.

Looking across timepoints in preterm infants, the SES measure most strongly associated with FA changed over time, from maternal education in the neonatal period to neighborhood deprivation at five years. This is consistent with previous work in this cohort showing that family-level SES (i.e. maternal education) was associated with more regional brain volumes than neighborhood-level SES (i.e. SIMD) at term-equivalent age (35), and findings that SIMD was associated with developmental outcomes in later infancy, including a preference to viewing social stimuli (71) and emotional regulation and cortisol responses (72). There is further evidence in the wider literature that neighborhood measures may matter more than parental education as children age (73–76). This may be due to family-level SES measures capturing the in utero and perinatal exposures, whereas neighborhood-level SES measures that capture wider socioeconomic circumstances become more influential over time.

### 4.2 Strengths and limitations

This is a rare cohort study of preterm and term children including MRI at two timepoints linked to detailed characterization of medical and social data. The preterm children did not have major parenchymal brain injuries, so are representative of most survivors of modern intensive care practices (21,22,41). We included two measures of SES, allowing comparison of how dimensions of SES vary in their association with FA. We adjusted for preterm exposures, testing the robustness of GA-FA and SES-FA associations in the context of other disease processes affecting brain dysmaturation.

FA is an established marker of encephalopathy of prematurity that is sensitive to upstream determinants of brain development and is associated with neurocognitive outcomes (7,77), and we used an established processing pipeline that is unbiased to particular tracts. There are some limitations to the use of FA as a metric of white matter microstructure; FA differences can represent differences in a number of different microstructural elements in a voxel (e.g. fiber density, myelination), but because of the complexity of the white matter with crossing fibers, FA differences could be due to disproportionate development or degeneration of a fiber bundle (78). However, there are studies showing association between FA and histological myelination (79), lending support to the use of FA as a proxy for white matter integrity.

There are some limitations. Although the cohort is comparable to neonatal populations in high-income, highly educated, majority-white settings, further work is needed to determine whether the social patterns we observed are generalizable to settings with different socioeconomic or ethnicity profiles. We studied two SES measures, but did not have data for all that could be relevant, such as household income, nor could we study the impact of social mobility on FA in this study group (80–82).

The 5-year sample is smaller than the neonatal sample because data collection at 5 years is ongoing. Nevertheless, based on previous sensitivity analyses of simulated data we estimate that in the smallest groupwise comparison we performed TBSS should identify differences in FA in 60-70% of affected voxels (83). Longitudinal analyses using dMRI metrics and behavioral testing are planned to better understand how SES and preterm birth might affect the rate of cerebral development and functional abilities over childhood (21,22).

### 4.3 Conclusions

Family- and neighborhood-level measures of SES are associated with white matter microstructure in the developing brain across childhood. However, the nature and direction of these associations are influenced by preterm birth and specific SES measures. In preterm infants, low GA and SES are associated with white matter microstructure in the newborn period and at five years, but the SES measure most closely associated with FA changes from maternal education in the neonatal period to neighborhood deprivation at five years. Preterm birth appears to expose a susceptibility of the developing brain to social disadvantage that persists through childhood. Strategies to mitigate the adverse effects of social disadvantage on child development may require a personalized approach that takes account of gestational age at birth and the timing of intervention, focusing on family-level interventions in very early life with increasing focus on neighborhood-level interventions as children grow up.

## Data Availability

All data produced in the present study are available upon reasonable request to the authors.

## Acknowledgements and funding

Funding to establish Theirworld Edinburgh Birth Cohort (TEBC) was from Theirworld (www.theirworld.org). MRI at five years is funded by the PRENCOG study (PReterm birth as a determinant of Neurodevelopment and COGnition in children), funded by a UKRI Medical Research Council Programme Grant, MR/X003434/1.

Participant MRI scans were carried out at the Edinburgh Imaging Facility (Royal Infirmary of Edinburgh), University of Edinburgh, which was established with funding from The Wellcome Trust, Dunhill Medical Trust, Edinburgh and Lothians Research Foundation, Theirworld, The Muir Maxwell Trust and other sources.

KM receives salary from NHS Lothian.

SA is funded by The German Academic Scholarship Foundation (Studienstiftung des deutschen Volkes).

RS is a student on the Translational Neuroscience PhD Programme, funded by Wellcome (218493/Z/19/Z).

GS is funded by an MRC Clinician Scientist Fellowship MR/X019535/1. CB is funded by an NIHR Advanced Fellowship (award ID NIHR300617).

GDB is supported by the UK Medical Research Council (MR/P023444/1) and the US National Institute on Aging (1R56AG052519-01, 1R01AG052519-01A1).

SRC is supported by a Sir Henry Dale Fellowship jointly funded by the Wellcome Trust and the Royal Society (221890/Z/20/Z).

MJT is funded by the NHS Lothian Research and Development Office.

The authors are grateful to the families who consented to take part in the study, and the Edinburgh Imaging Team for providing infant and child scanning.

This article was published as a preprint on medRxiv.

## CRediT author contributions

KM: conceptualization, methodology, formal analysis, investigation, resources, data curation, writing (original draft), writing (review/editing), visualization.

MBC: conceptualization, methodology, software, formal analysis, investigation, data curation, writing (review/editing), visualization.

MT: conceptualization, methodology, software, formal analysis, investigation, data curation, writing (review/editing), visualization.

SA: investigation, resources, writing (review/editing). RS: investigation, writing (review/editing).

JS: investigation, writing (review/editing).

LJS: investigation, resources, writing (review/editing).

KV: formal analysis, investigation, writing (review/editing).

GS: investigation, writing (review/editing).

JH: data curation, project administration, writing (review/editing).

AC: data curation, project administration, writing (review/editing).

GB: investigation, resources, writing (reviewing/editing).

CJ: investigation, resources, writing (reviewing/editing).

IG: investigation, resources, writing (reviewing/editing).

DM: investigation, resources, writing (reviewing/editing).

YWC: investigation, resources, data curation, writing (review/editing).

RA: investigation, resources, data curation, writing (review/editing).

AJQ: investigation, resources, writing (review/editing).

CB: writing (review/editing), funding acquisition.

AT: writing (review/editing), funding acquisition.

GDB: writing (review/editing), funding acquisition.

RMR: writing (review/editing), funding acquisition.

SRC: methodology, writing (review/editing), funding acquisition.

HCW: methodology, writing (review/editing), funding acquisition.

MJT: methodology, resources, writing (review/editing).

MEB: software, resources, writing (review/editing), funding acquisition.

HR: conceptualization, methodology, formal analysis, resources, writing (original draft), writing (review/editing), visualization, supervision, project administration, funding acquisition.

JPB: conceptualization, methodology, formal analysis, resources, writing (original draft), writing (review/editing), visualization, supervision, project administration, funding acquisition.

