## Supplement for "Preterm birth, socioeconomic status, and white matter development across childhood"

**Contents**

Supplementary eMethods

Figure S1. Flowchart of included participants

Supplementary analyses – whole cohort

- Table S1. TBSS multivariable linear regression models for whole cohort analyses
- Figure S2. Voxels where FA associations were found in whole cohort analyses
- Figure S3. Interaction effect between GA and SIMD on FA at the neonatal timepoint in the whole cohort
- Figure S4. Interaction effect between GA and SIMD on FA at five years of age in the whole cohort
- Table S2. Number of significant voxels where FA associates with GA, SES, or an interaction between GA and SES for the whole cohort

Table S3. TBSS multivariable linear regression models

Table S4. Change in the number of voxels associated with FA after adjustment for co-morbidities of preterm birth

Figure S5. Interaction effects between GA and SIMD on FA at five years of age in preterm children

Supplementary references

**Supplementary eMethods**

*Longer MRI scanning protocol*

- Neonatal timepoint: three-dimensional (3D) T1-weighted magnetization-prepared rapid acquisition with gradient echo (MPRAGE) structural volume scan, 3D T2-weighted sampling perfection with application-optimized contrasts by using flip angle evolution (SPACE) structural scan, 3D susceptibility-weighted imaging, axial 2D fluid-attenuated inversion-recovery (FLAIR) imaging, and magnetization transfer saturation (MTSat) imaging.
- Five-year timepoint: 3D T2-weighted SPACE, FLAIR, and MTSat imaging, and functional MRI using naturalistic child-friendly film stimuli (Pixar's "Partly Cloudy" and clips from Sesame Workshop's "Sesame Street").

*Maternal education levels*

- None: no qualifications obtained.
- Basic high school qualification: National 5s, Standard Grades, GCSEs (General Certificates of Secondary Education) or equivalent. This is divided into 1-4 and >4 qualifications.
- Advanced high school qualification: Highers, A levels (Advanced levels) or equivalent.
- College qualification: e.g. National Certificate, Higher National Diploma, Higher National Certificate, vocational qualifications.
- University undergraduate degree.
- University postgraduate degree.

*Preterm exposure definitions*

- Sepsis: positive blood culture and/or physician decision to treat with five days of antibiotics, including both early- and late-onset sepsis.
- Histological chorioamnionitis (HCA): confirmed by histological analysis by histopathologist.
- Bronchopulmonary dysplasia (BPD): requirement for respiratory support and/or supplemental oxygen after 36 weeks' corrected gestation.
- Necrotizing enterocolitis (NEC): includes both medical (7 days nil by mouth) or surgical management.
- Low breast milk intake: defined as <75% days during neonatal unit stay with exclusive maternal or donor breast milk.

These measures were gathered from medical records, and counted as binary yes/no for presence, with a total score 0-5 (1,2), and were only assessed in preterm children.

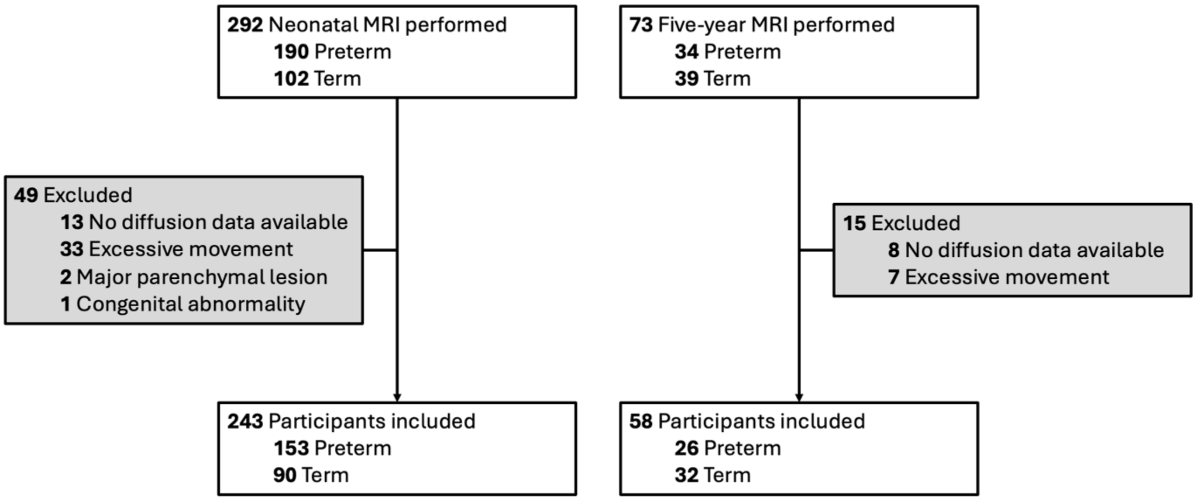


**Figure S1. Flowchart of included participants**

Neonatal MRI scanning took place between April 2017 and December 2021. Five-year MRI scanning took place between April 2022 and February 2024.

**Supplementary analyses – whole cohort**

In pre-registered analyses, to maximize our sample size, we performed analyses of the whole cohort at each timepoint (neonatal timepoint n=243, five-year timepoint n=58). The tract-based spatial statistics (TBSS) general linear regression models are detailed in Table S2.

**Table S1. TBSS multivariable linear regression models for whole cohort analyses.**

| Timepoint | Model |
|---|---|
| Neonatal | Neonatal FA ~ GA + maternal education + GA*maternal education + sex + age at MRI<br>Neonatal FA ~ GA + SIMD + GA*SIMD + sex + age at MRI |
| Five-year | Five-year FA ~ GA + maternal education + GA*maternal education + sex + age at MRI<br>Five-year FA ~ GA + SIMD + GA*SIMD + sex + age at MRI |

An intercept was also included. If no multiplication interaction effect was seen, this was removed.

*Neonatal timepoint: whole cohort results*

In the whole cohort (n=243), GA associated with FA across the white matter skeleton (Figure S2A). Correlations were primarily positive, in that higher GA corresponded to higher FA values, but there were also areas with negative correlations in the cerebellum and brainstem. The pattern of results was similar regardless of the SES measure included in the model. Figure S2A shows results adjusted for maternal education and similar results were seen using SIMD.

In contrast, an interaction between GA and maternal education associated with lower FA in smaller regions of the external capsule bilaterally (Figure S2B). This interaction effect describes a negative association between higher birth GA and higher FA for infants of mothers with no educational qualifications (Figure S3); this includes only 5 children, all born preterm ($\beta$=-0.004, $p$=0.008). For infants of mothers with any educational qualifications, from basic high school qualifications to university postgraduate qualifications, there was a positive association between higher birth GA and higher FA ($\beta$ range=0.0007-0.004, $p$=0.008).

There were no associations between maternal education alone with FA, nor were there SIMD or SIMD×GA associations with FA.

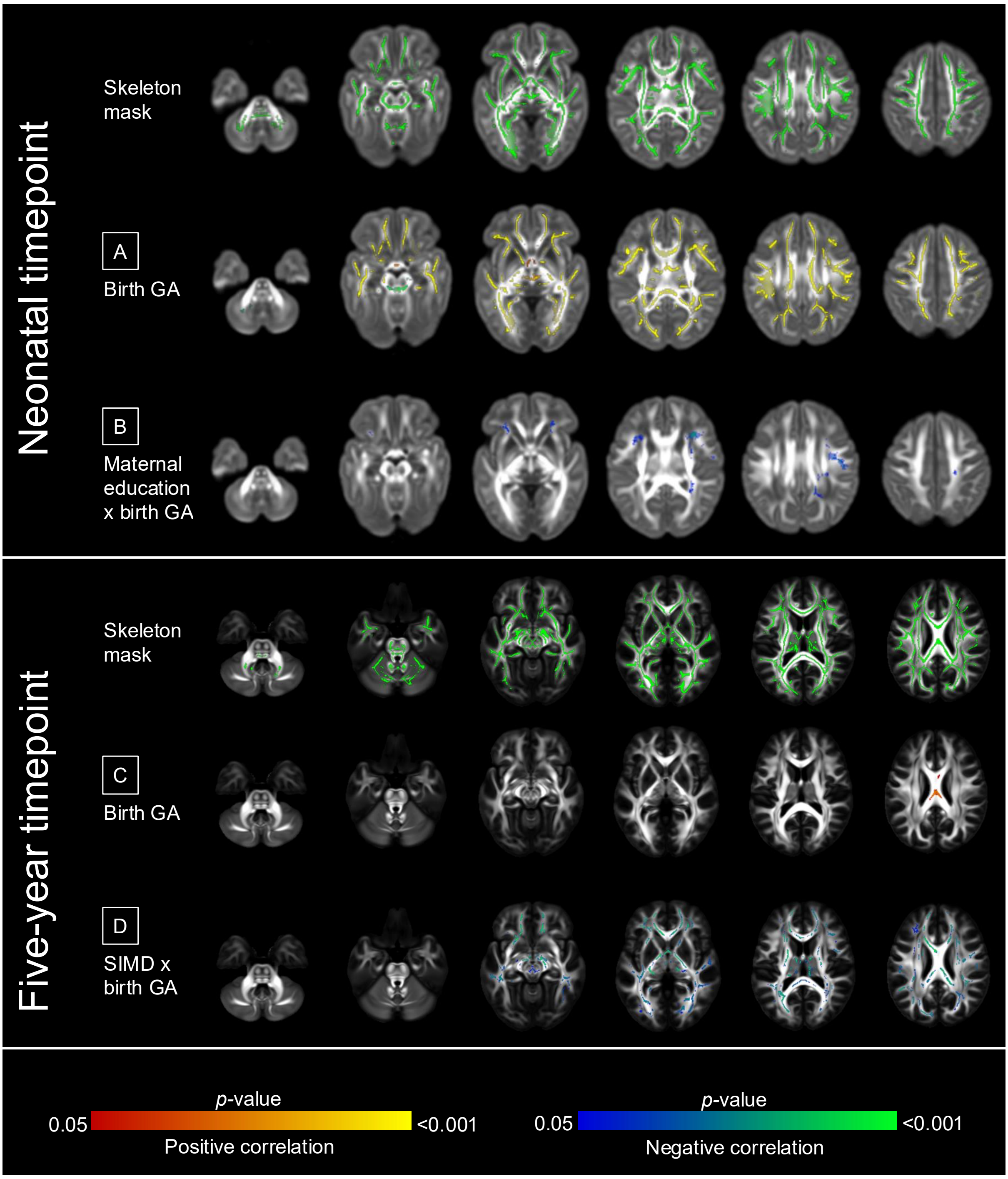


**Figure S2. Voxels where FA associations were found in whole cohort analyses.**

Voxels where FA associations were found in the whole cohort at the neonatal timepoint (n=243) or at five years of age (n=58). Models were mutually adjusted for GA, SES (maternal education or SIMD), sex, and age at MRI. At the neonatal timepoint – A: GA, B: Maternal education × GA. At five years – C: GA, D: SIMD × GA.

Results are reported after 5000 permutations, p-values corrected using threshold-free cluster enhancement and family-wise error correction with a significance level of $p<0.05$. For visualization: anatomic left is on the left side of the image. Red-yellow indicates a positive association, blue indicates a negative association. Overlaid on the developing Human Connectome Project (dHCP) neonatal template (3) for the neonatal timepoint, and on the FSL HCP1065 (Human Connectome Project) adult template (4) for the five-year timepoint.

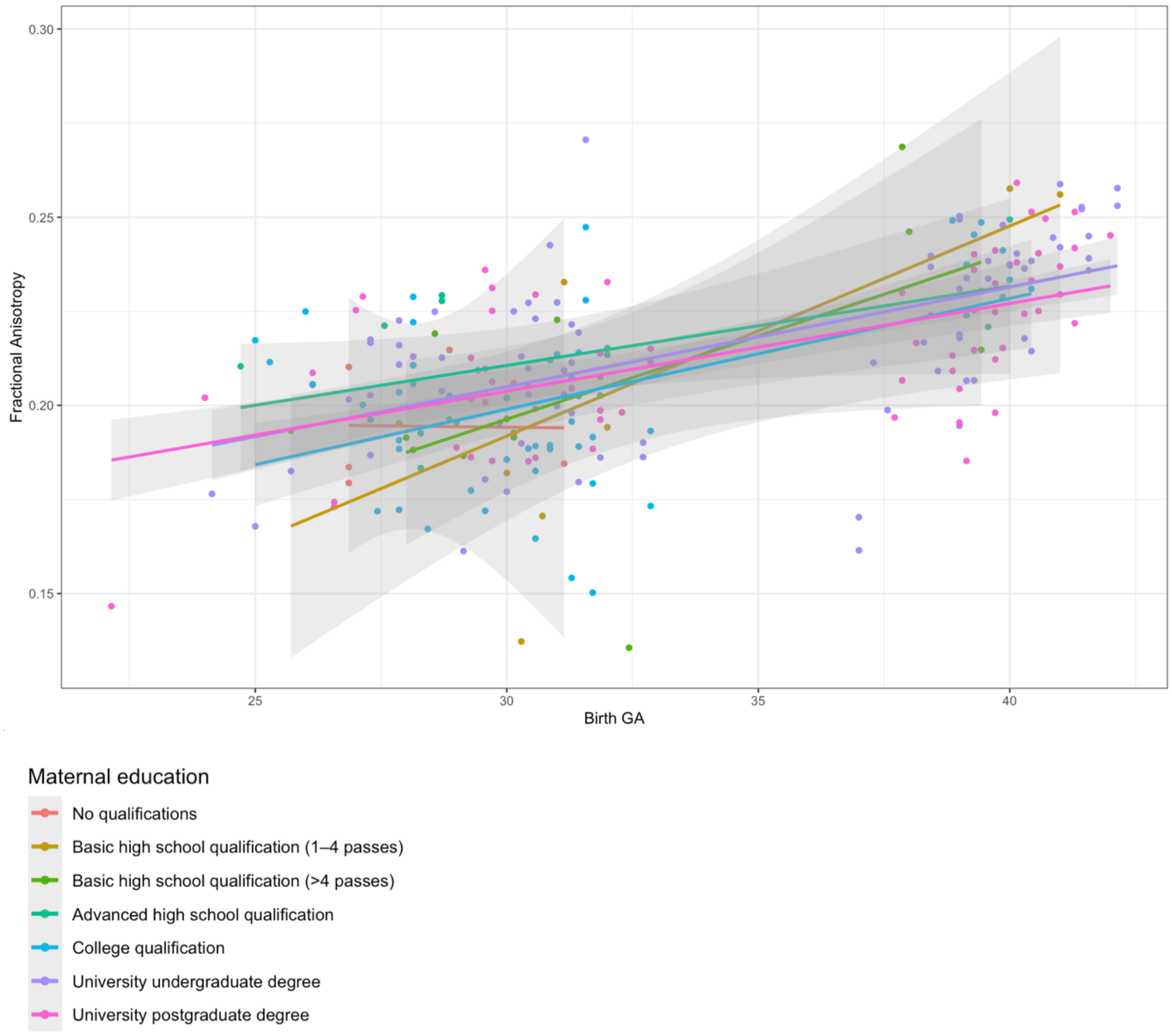


**Figure S3. Interaction effect between GA and maternal education on FA at the neonatal timepoint in the whole cohort**

Interaction effect between GA and maternal education on FA in the whole cohort (n=243). For each individual, mean FA was extracted from voxels where there are significant interaction effects. Maternal education analyses were performed using maternal education as seven categories.

*Five-year timepoint: whole cohort results*

In the whole cohort (n=58), in comparison to the widespread associations between FA and GA at the neonatal timepoint, there were only small regions where higher GA associated with higher FA, in the corpus callosum.

There were anatomically widespread interaction effects between GA and SIMD on FA. To illustrate this interaction, this is visualized using SIMD quintile 1-4 (n=36) and quintile 5 (n=22) (Figure S4). This interaction was such that higher GA associated with higher FA for children from SIMD 1-4 (i.e. whose families are living in more deprived neighborhoods), but higher GA associated with lower FA for children from SIMD 5 (i.e. whose families are living in the least deprived neighborhoods) ($\beta$=0.004 and $\beta$=-0.002 respectively, $p$<0.001). There were no associations with SIMD or maternal education independently, or interaction effects between maternal education and GA on FA.

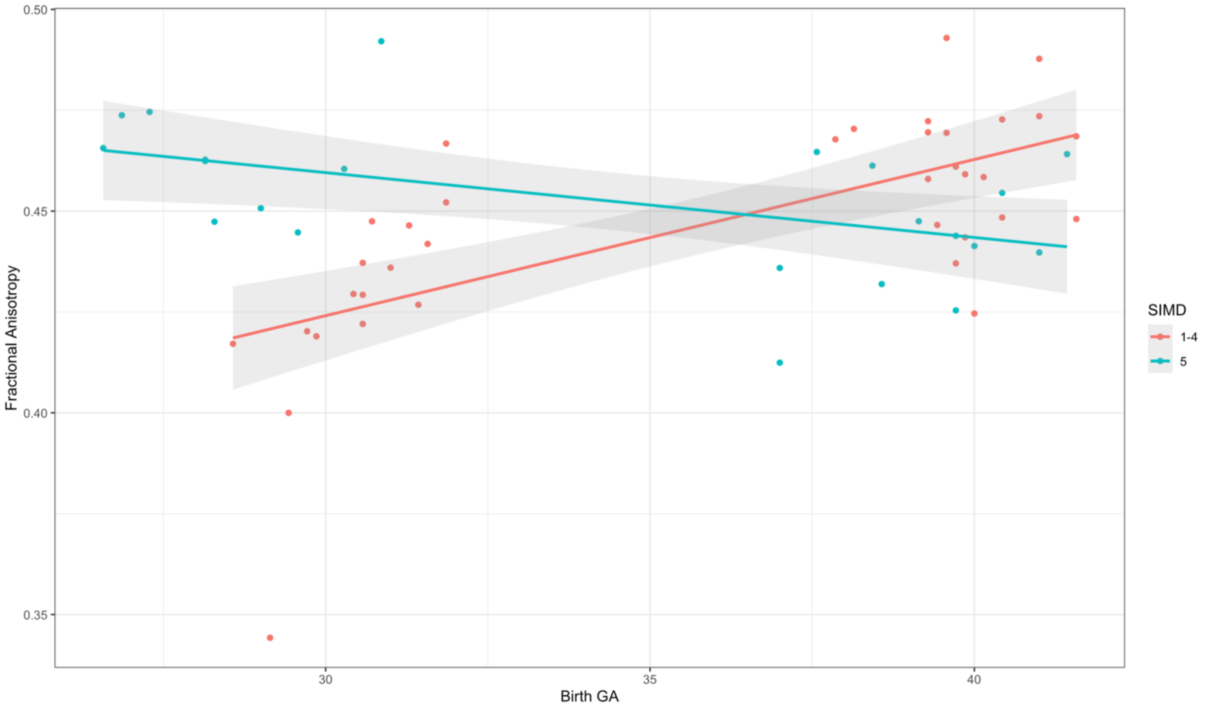


**Figure S4. Interaction effect between GA and SIMD on FA at five years of age in the whole cohort.**

Interaction effect between GA and SIMD on FA at five years of age in the whole cohort (n=58). For each individual, mean FA was extracted from voxels where there are significant interaction effects. Analyses were performed with SIMD rank as a continuous measure. It is visualized here as linear regression lines for two groups for ease of interpretation, namely SIMD quintiles 1-4 and SIMD quintile 5.

*Across timepoint results*

In the whole cohort, there was a difference in the proportions of the white matter skeleton with SES×GA-FA interaction associations between the neonatal period and five years ($p$<0.001), but not for SES-FA associations.

At the neonatal timepoint, FA was associated with a multiplication interaction between maternal education and GA (5.2%, 1541/29354), but not SIMD×GA. At the five-year timepoint, FA was associated with an interaction between GA and SIMD (30.2%, 31434/104030), but not maternal education×GA. There were no independent associations between maternal education or SES and FA at either timepoint. Details of the number and proportion of significant voxels where there were associations between FA and GA, SES, SES×GA for the whole cohort are in Table S3.

**Table S2. Number of significant voxels where FA associates with GA, SES, or an interaction between GA and SES for the whole cohort**

| Timepoint | Variable | Number (proportion) of significant voxels |
|---|---|---|
| Neonatal | GA (in model with maternal education) | 23275 (79.3%) |
| | GA (in model with SIMD) | 23759 (80.9%) |
| | Maternal education | 0 |
| | Maternal education × GA | 1541 (5.2%) |
| | SIMD | 0 |
| | SIMD x GA | 0 |
| Five-year | GA (in model with maternal education) | 528 (0.5%) |
| | GA (in model with SIMD) | 635 (0.6%) |
| | Maternal education | 0 |
| | Maternal education × GA | 0 |
| | SIMD | 0 |
| | SIMD × GA | 31434 (30.2%) |

Number of significant voxels ($p_{FWER}$<0.05) where FA associates with GA, SES, or an interaction between GA and SES. Total number of voxels at neonatal timepoint = 29354, and at five years of age = 104030.

**Table S3. TBSS multivariable linear regression models.**

| Timepoint | Group | Model |
|---|---|---|
| Neonatal | Preterm | Neonatal FA ~ GA + maternal education + GA*maternal education + sex + age at MRI |
| | | Neonatal FA ~ GA + SIMD + GA*SIMD + sex + age at MRI |
| | | Neonatal FA ~ GA + maternal education + GA*maternal education + sex + age at MRI + preterm exposures |
| | | Neonatal FA ~ GA + SIMD + GA*SIMD + sex + age at MRI + preterm exposures |
| | Term | Neonatal FA ~ GA + maternal education + GA*maternal education + sex + age at MRI |
| | | Neonatal FA ~ GA + SIMD + GA*SIMD + sex + age at MRI |
| Five-year | Preterm | Five-year FA ~ GA + maternal education + GA*maternal education + sex + age at MRI |
| | | Five-year FA ~ GA + SIMD + GA*SIMD + sex + age at MRI |
| | | Five-year FA ~ GA + maternal education + GA*maternal education + sex + age at MRI + preterm exposures |
| | | Five-year FA ~ GA + SIMD + GA*SIMD + sex + age at MRI + preterm exposures |
| | Term | Five-year FA ~ GA + maternal education + GA*maternal education + sex + age at MRI |
| | | Five-year FA ~ GA + SIMD + GA*SIMD + sex + age at MRI |

An intercept was also included. If no multiplication interaction effect was seen, this was removed.

Preterm exposures is the sum of sepsis, HCA, BPD, NEC, and low breast milk intake.

**Table S4. Change in the number of voxels associated with FA after adjustment for co-morbidities of preterm birth**

| Timepoint | Variable | Number (proportion) of significant voxels in unadjusted model | Number (proportion) of significant voxels in adjusted model |
|---|---|---|---|
| Neonatal | GA (in model with maternal education) | 3934 (13.4%) | 0 |
| | GA (in model with SIMD) | 4014 (13.7%) | 0 |
| | Maternal education | 16890 (57.5%) | 15949 (54.3%) |
| | Maternal education × GA | 15544 (53%) | 16175 (55.1%) |
| | SIMD | 0 | 0 |
| | SIMD × GA | 2526 (8.6%) | 7001 (23.9%) |
| Five-year | GA (in model with maternal education) | 0 | 0 |
| | GA (in model with SIMD) | 5816 (5.6%) | 959 (0.9%) |
| | Maternal education | 0 | 0 |
| | Maternal education × GA | 0 | 0 |
| | SIMD | 15381 (14.8%) | 21281 (20.5%) |
| | SIMD × GA | 7008 (6.7%) | 7166 (6.9%) |

Change in significant voxels in the preterm infants where FA associates with GA, SES, or an interaction between GA and SES, before and after adjustment for preterm factors (BPD, HCA, low breast milk, NEC, and sepsis).

Total number of voxels at neonatal timepoint = 29354, and at five years of age = 104030.

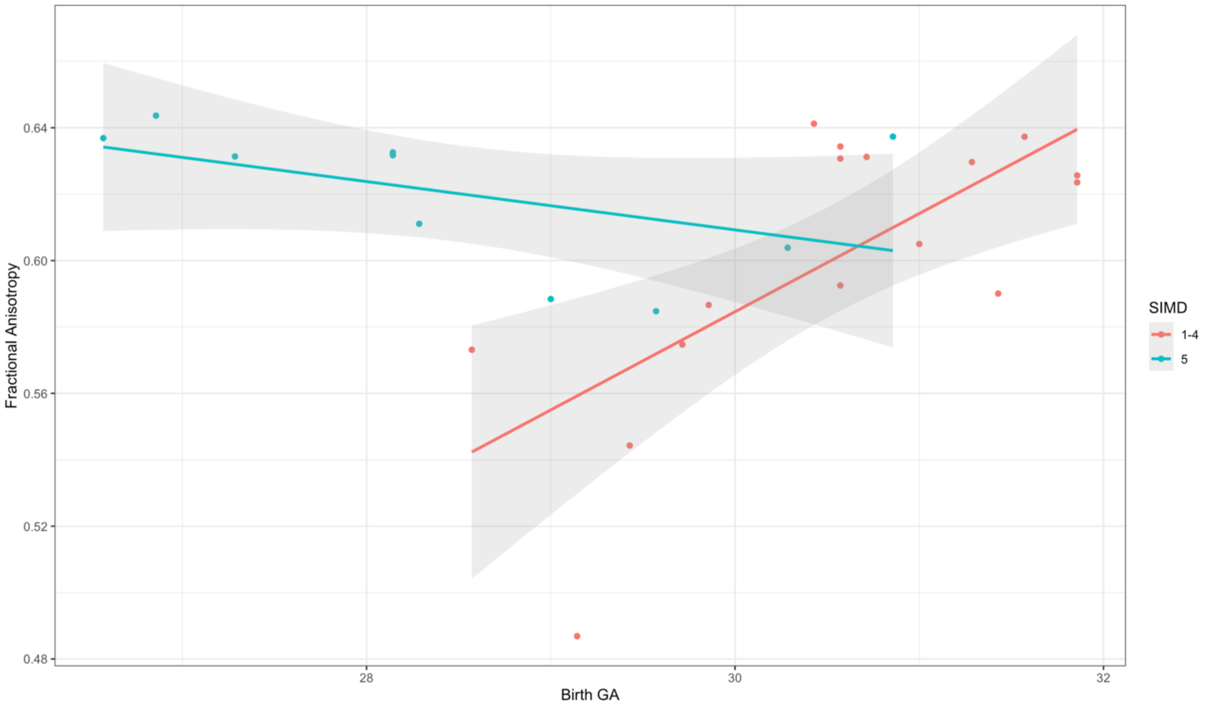


**Figure S5. Interaction effects between GA and SIMD on FA at five years of age in preterm children.**

Interaction effect between GA and SIMD on FA at five years of age in preterm children (n=26). For each individual, mean FA was extracted from voxels where there are significant interaction effects. Analyses were performed with SIMD rank as a continuous measure. It is visualized here as linear regression lines for two groups for ease of interpretation, namely SIMD quintiles 1-4 and SIMD quintile 5.
